# Duration of immune protection of SARS-CoV-2 natural infection against reinfection in Qatar

**DOI:** 10.1101/2022.07.06.22277306

**Authors:** Hiam Chemaitelly, Nico Nagelkerke, Houssein H. Ayoub, Peter Coyle, Patrick Tang, Hadi M. Yassine, Hebah A. Al-Khatib, Maria K. Smatti, Mohammad R. Hasan, Zaina Al-Kanaani, Einas Al-Kuwari, Andrew Jeremijenko, Anvar Hassan Kaleeckal, Ali Nizar Latif, Riyazuddin Mohammad Shaik, Hanan F. Abdul-Rahim, Gheyath K. Nasrallah, Mohamed Ghaith Al-Kuwari, Adeel A. Butt, Hamad Eid Al-Romaihi, Mohamed H. Al-Thani, Abdullatif Al-Khal, Roberto Bertollini, Laith J. Abu-Raddad

## Abstract

**BACKGROUND:** The future of the SARS-CoV-2 pandemic hinges on virus evolution and duration of immune protection of natural infection against reinfection. We investigated duration of protection afforded by natural infection, the effect of viral immune evasion on duration of protection, and protection against severe reinfection, in Qatar, between February 28, 2020 and June 5, 2022.

**METHODS:** Three national, matched, retrospective cohort studies were conducted to compare incidence of SARS-CoV-2 infection and COVID-19 severity among unvaccinated persons with a documented SARS-CoV-2 primary infection, to incidence among those infection-naïve and unvaccinated. Associations were estimated using Cox proportional-hazard regression models.

**RESULTS:** Effectiveness of pre-Omicron primary infection against pre-Omicron reinfection was 85.5% (95% CI: 84.8-86.2%). Effectiveness peaked at 90.5% (95% CI: 88.4-92.3%) in the 7^th^ month after the primary infection, but waned to ∼70% by the 16^th^ month. Extrapolating this waning trend using a Gompertz curve suggested an effectiveness of 50% in the 22^nd^ month and <10% by the 32^nd^ month. Effectiveness of pre-Omicron primary infection against Omicron reinfection was 38.1% (95% CI: 36.3-39.8%) and declined with time since primary infection. A Gompertz curve suggested an effectiveness of <10% by the 15^th^ month. Effectiveness of primary infection against severe, critical, or fatal COVID-19 reinfection was 97.3% (95% CI: 94.9- 98.6%), irrespective of the variant of primary infection or reinfection, and with no evidence for waning. Similar results were found in sub-group analyses for those ≥50 years of age.

**CONCLUSIONS:** Protection of natural infection against reinfection wanes and may diminish within a few years. Viral immune evasion accelerates this waning. Protection against severe reinfection remains very strong, with no evidence for waning, irrespective of variant, for over 14 months after primary infection.

## Introduction

The future of the severe acute respiratory syndrome coronavirus 2 (SARS-CoV-2) pandemic is uncertain, but it hinges on virus evolution and the level and duration of immune protection of natural infection against reinfection.^1–3^ While current coronavirus disease 2019 (COVID-19) vaccines had a critical role in reducing COVID-19 hospitalizations and deaths, their rapidly waning immune protection, particularly against the Omicron (B.1.1.529) variant,^4–8^ limits their role in shaping the future of SARS-CoV-2 epidemiology compared to other vaccines, such as vaccinia, which eradicated smallpox.^9^

Seasonal “common-cold” coronaviruses are characterized by short-term immunity against mild reinfection,^10^ but long-term immunity against severe reinfection.^2^ While SARS-CoV-2 infection with the original virus or pre-Omicron variants elicited >80% protection against reinfection with the original virus^11–13^ or with Alpha (B.1.1.7),^14^ Beta (B.1.351),^15^ and Delta (B.1.617.2)^16^ variants, protection against reinfection with Omicron subvariants is below 60%.^16, 17^ Reinfections have become common since Omicron emergence.^17^

We sought to answer three questions of relevance to the future of this pandemic: 1) When infected with a pre-Omicron variant, how long does protection persist against reinfection with pre-Omicron variants? 2) When infected with a pre-Omicron variant, how long does protection persist against reinfection with an Omicron subvariant? 3) When infected with any variant, how long does protection persist against severe, critical, or fatal COVID-19? Answers to these questions help us to understand duration of protection resulting from natural-infection, effects of viral evasion of the immune system on this duration, and effectiveness of natural infection against COVID-19 severity when reinfection occurs.

Three studies were conducted to answer these questions in Qatar, a country that experienced five SARS-CoV-2 waves dominated by each of the original virus,^11^ Alpha,^14^ Beta,^15^ Omicron BA.1 and BA.2,^18^ and currently Omicron BA.4 and BA.5, in addition to a prolonged low-incidence phase dominated by Delta.^4^

## Methods

### Study population and data sources

This study was conducted in the population of Qatar and analyzed COVID-19 data for laboratory testing, vaccination, hospitalization, and death, retrieved from the national digital-health information platform. Databases include all SARS-CoV-2-related data, with no missing information since pandemic onset, such as all polymerase chain reaction (PCR) tests, and starting from January 5, 2022, rapid antigen tests conducted at healthcare facilities. Further descriptions of the study population and these national databases were reported previously.^4, 15, 17, 19, 20^

### Study designs and cohorts

We conducted three matched, retrospective cohort studies that emulated randomized “target” trials.^20, 21^ In each study, incidence of infection or of severe,^22^ critical,^22^ or fatal^23^ COVID-19 was compared in the national cohort of individuals with a documented SARS-CoV-2 primary (first) infection prior to vaccination (designated the primary-infection cohort) to the national (control) cohort of individuals who are infection-naïve and unvaccinated (designated the infection-naïve cohort).

Documentation of infection in all cohorts was based on positive PCR or rapid antigen tests. Laboratory methods are found in Supplementary Appendix Section S1. Classification of COVID-19 case severity (acute-care hospitalizations),^22^ criticality (intensive-care-unit hospitalizations),^22^ and fatality^23^ followed World Health Organization guidelines (Section S2).

### Cohort matching and follow-up

Individuals in the primary-infection cohort were exact-matched in a one-to-one ratio by sex, 10- year age group, nationality, and comorbidity count (none, 1-2 comorbidities, 3 or more comorbidities) to individuals in the infection-naïve cohort, to control for differences in risk of SARS-CoV-2 infection in Qatar.^19, 24–27^ Matching by these factors was shown to provide adequate control of differences in risk of infection.^4, 28–31^ Matching was also done by the calendar week of SARS-CoV-2 testing. That is, an individual who was diagnosed with a primary infection in a specific calendar week was matched to an infection-naïve individual who had a record of a SARS-CoV-2-negative test in that same week (Figures S1-S3). This matching ensures that all individuals in all cohorts had active presence in Qatar at the same calendar time. Matching was performed using an iterative process so that each individual in the infection-naïve cohort was alive, infection-free, and unvaccinated at the start of follow-up.

SARS-CoV-2 reinfection is conventionally defined as a documented infection ≥90 days after an earlier infection, to avoid misclassification of prolonged PCR positivity as reinfection.^12, 13, 16^ Therefore, each matched pair was followed from the calendar day an individual in the primary-infection cohort completed 90 days after a documented primary infection.

For exchangeability, both members of each matched pair were censored on the date of first-dose vaccination of an individual in either cohort.^20, 32^ Individuals were followed up until the first of any of the following events: a documented SARS-CoV-2 infection, i.e., the first PCR-positive or rapid-antigen-positive test after the start of follow-up, regardless of symptoms, or first-dose vaccination (with matched pair censoring), or death, or end of study censoring.

### Pre-Omicron Reinfection Study

This study estimated effectiveness of a pre-Omicron primary infection against reinfection with a pre-Omicron variant. Any individual with a documented primary infection between February 28, 2020 (pandemic onset in Qatar) and November 30, 2021 was eligible for inclusion in the primary-infection cohort, provided that the individual received no vaccination before the start of follow-up, 90 days after primary infection. Any individual with a SARS-CoV-2-negative test during this period was eligible for inclusion in the infection-naïve cohort, provided that the individual had no record of infection or vaccination before the start of follow-up. Follow-up was from the 90^th^ day after primary infection until November 30, 2021 (first evidence of Omicron in Qatar^16, 20^ to ensure that incidence during the study was only due to a pre-Omicron variant). The primary study outcome was incidence of infection. The secondary outcome was incidence of severe, critical, or fatal COVID-19.

### Omicron Reinfection Study

This study estimated effectiveness of a pre-Omicron primary infection against reinfection with an Omicron subvariant. Any individual with a documented primary infection from February 28, 2020 until November 30, 2021 was eligible for inclusion in the primary-infection cohort, absent any record of reinfection or vaccination before the start of follow-up. Any individual with a SARS-CoV-2-negative test during this period was eligible for inclusion in the infection-naïve cohort, absent any record of infection or vaccination before the start of follow-up. Follow-up was from December 19, 2021 (onset of the Omicron wave in Qatar),^16, 20^ if the primary infection occurred ≥90 days before this date. Follow-up was from the 90^th^ day after primary infection if the primary infection occurred <90 days before December 19, 2021. Follow-up was until June 5, 2022. The primary study outcome was incidence of infection. The secondary outcome was incidence of severe, critical, or fatal COVID-19.

### COVID-19 Severity Reinfection Study

This study estimated effectiveness of a primary infection against severe, critical, or fatal COVID-19 reinfection, irrespective of the variant of primary infection or reinfection. Any individual with a documented primary infection between February 28, 2020 and June 5, 2022 was eligible for inclusion in the primary-infection cohort, provided that the individual received no vaccination before the start of follow-up, 90 days after primary infection. Any individual with a SARS-CoV-2-negative test during this period was eligible for inclusion in the infection-naïve cohort, absent any record of infection or vaccination before the start of follow-up. The primary study outcome was incidence of severe, critical, or fatal COVID-19. The secondary outcome was incidence of infection.

### Statistical analysis

Eligible and matched cohorts were described using frequency distributions and measures of central tendency, and were compared using standardized mean differences (SMDs). An SMD <0.1 indicated adequate matching. Cumulative incidence of infection (defined as the proportion of individuals at risk, whose primary endpoint during follow-up was a reinfection for the primary-infection cohort, or an infection for the infection-naïve cohort) was estimated using the Kaplan–Meier estimator method. Incidence rate of infection in each cohort, defined as the number of identified infections divided by the number of person-weeks contributed by all individuals in the cohort, was estimated with its 95% confidence interval (CI) using a Poisson log-likelihood regression model with the Stata 17.0 *stptime* command.

The hazard ratio, comparing incidence of infection in both cohorts, and the corresponding 95% CI were calculated using Cox regression adjusted for matching factors with the Stata 17.0 *stcox* command. Schoenfeld residuals and log-log plots for survival curves were used to test the proportional-hazards assumption and to investigate its adequacy. 95% CIs were not adjusted for multiplicity; thus, they should not be used to infer definitive differences between cohorts.

Interactions were not considered. Effectiveness against reinfection was estimated using the equation: Effectiveness = 1−adjusted hazard ratio . Analogous analyses were used when the outcome was severe, critical, or fatal COVID-19.

Subgroup analyses were conducted to investigate waning of protection over time. Adjusted hazard ratios were estimated by month since primary infection using separate Cox regressions with "failures" restricted to specific months, in the Pre-Omicron Reinfection Study and the COVID-19 Severity Reinfection Study. Adjusted hazard ratios were also calculated in the Omicron Reinfection Study, but stratified by 3-calendar-month primary-infection sub-cohorts. Additional analyses restricting matched cohorts to those ≥50 years of age were conducted.

Sensitivity analyses adjusting effectiveness estimates for differences in testing frequency between cohorts were conducted.

Waning of protection was fitted to the Gompertz function^33^ using the Stata 17.0 *nl* command. This function has been used to describe decay of immunity, such as against smallpox,^33^ and provided a suitable description of waning of protection as assessed by an empiric goodness-of-fit. Statistical analyses were conducted using Stata/SE version 17.0 (Stata Corporation, College Station, TX, USA).

### Oversight

Hamad Medical Corporation and Weill Cornell Medicine-Qatar Institutional Review Boards approved this retrospective study with a waiver of informed consent. The study was reported following the Strengthening the Reporting of Observational Studies in Epidemiology (STROBE) guidelines. The STROBE checklist is found in Table S1.

## Results

### Pre-Omicron Reinfection Study

Figure S1 shows the population selection process and Table 1 describes baseline characteristics of the full and matched cohorts. The matched cohorts each included 290,638 individuals. The study was conducted on the total population of Qatar, and thus the study population is representative of the internationally diverse, but predominantly young and male population of Qatar (Table S2).

**Table 1.** Baseline characteristics of the eligible and matched primary-infection and infection-naïve cohorts in the Pre-Omicron Reinfection Study and the Omicron Reinfection Study.

| Characteristics | Pre-Omicron Reinfection Study |  |  |  |  |  | Omicron Reinfection Study |  |  |  |  |  |
| --- | --- | --- | --- | --- | --- | --- | --- | --- | --- | --- | --- | --- |
|  | Full eligible cohorts |  |  | Matched cohorts* |  |  | Full eligible cohorts |  |  | Matched cohorts* |  |  |
|  | Primary-infection cohort | Infection-naïve cohort | SMD† | Primary-infection cohort | Infection-naïve cohort | SMD† | Primary-infection cohort | Infection-naïve cohort | SMD† | Primary-infection cohort | Infection-naïve cohort | SMD† |
|  | N=301,943 | N=2,329,039 |  | N=290,638 | N=290,638 |  | N=127,075 | N=2,329,039 |  | N=120,483 | N=120,483 |  |
| Median age (IQR)—years | 32 (24-40) | 31 (24-39) | 0.05‡ | 32 (24-40) | 32 (24-40) | 0.00‡ | 27 (9-36) | 31 (24-39) | 0.36‡ | 27 (9-36) | 27 (9-36) | 0.00‡ |
| Age group |  |  |  |  |  |  |  |  |  |  |  |  |
| 0-9 years | 29,583 (9.8) | 258,890 (11.1) | 0.07 | 29,022 (10.0) | 29,022 (10.0) | 0.00 | 32,465 (25.6) | 258,890 (11.1) | 0.40 | 31,834 (26.4) | 31,834 (26.4) | 0.00 |
| 10-19 years | 22,830 (7.6) | 168,154 (7.2) |  | 21,900 (7.5) | 21,900 (7.5) |  | 11,338 (8.9) | 168,154 (7.2) |  | 10,630 (8.8) | 10,630 (8.8) |  |
| 20-29 years | 71,904 (23.8) | 585,421 (25.1) |  | 70,011 (24.1) | 70,011 (24.1) |  | 27,556 (21.7) | 585,421 (25.1) |  | 26,467 (22.0) | 26,467 (22.0) |  |
| 30-39 years | 96,468 (32.0) | 736,856 (31.6) |  | 93,754 (32.3) | 93,754 (32.3) |  | 32,736 (25.8) | 736,856 (31.6) |  | 31,340 (26.0) | 31,340 (26.0) |  |
| 40-49 years | 52,822 (17.5) | 363,820 (15.6) |  | 50,356 (17.3) | 50,356 (17.3) |  | 15,008 (11.8) | 363,820 (15.6) |  | 13,908 (11.5) | 13,908 (11.5) |  |
| 50-59 years | 20,772 (6.9) | 151,131 (6.5) |  | 19,234 (6.6) | 19,234 (6.6) |  | 5,453 (4.3) | 151,131 (6.5) |  | 4,597 (3.8) | 4,597 (3.8) |  |
| 60-69 years | 5,950 (2.0) | 50,033 (2.2) |  | 5,127 (1.8) | 5,127 (1.8) |  | 1,937 (1.5) | 50,033 (2.2) |  | 1,376 (1.1) | 1,376 (1.1) |  |
| 70+ years | 1,614 (0.5) | 14,734 (0.6) |  | 1,234 (0.4) | 1,234 (0.4) |  | 582 (0.5) | 14,734 (0.6) |  | 331 (0.3) | 331 (0.3) |  |
| Sex |  |  |  |  |  |  |  |  |  |  |  |  |
| Male | 220,978 (73.2) | 1,625,533 (69.8) | 0.08 | 212,685 (73.2) | 212,685 (73.2) | 0.00 | 85,134 (67.0) | 1,625,533 (69.8) | 0.06 | 80,791 (67.1) | 80,791 (67.1) | 0.00 |
| Female | 80,965 (26.8) | 703,506 (30.2) |  | 77,953 (26.8) | 77,953 (26.8) |  | 41,941 (33.0) | 703,506 (30.2) |  | 39,692 (32.9) | 39,692 (32.9) |  |
| Nationality§ |  |  |  |  |  |  |  |  |  |  |  |  |
| Bangladeshi | 26,984 (8.9) | 152,627 (6.6) | 0.24 | 25,217 (8.7) | 25,217 (8.7) | 0.00 | 5,808 (4.6) | 152,627 (6.6) | 0.19 | 5,280 (4.4) | 5,280 (4.4) | 0.00 |
| Egyptian | 15,517 (5.1) | 121,086 (5.2) |  | 15,275 (5.3) | 15,275 (5.3) |  | 7,789 (6.1) | 121,086 (5.2) |  | 7,525 (6.3) | 7,525 (6.3) |  |
| Filipino | 23,569 (7.8) | 169,647 (7.3) |  | 23,252 (8.0) | 23,252 (8.0) |  | 9,244 (7.3) | 169,647 (7.3) |  | 9,070 (7.5) | 9,070 (7.5) |  |
| Indian | 80,738 (26.7) | 641,424 (27.5) |  | 80,426 (27.7) | 80,426 (27.7) |  | 33,697 (26.5) | 641,424 (27.5) |  | 33,122 (27.5) | 33,122 (27.5) |  |
| Nepalese | 36,149 (12.0) | 201,681 (8.7) |  | 33,226 (11.4) | 33,226 (11.4) |  | 11,459 (9.0) | 201,681 (8.7) |  | 10,373 (8.6) | 10,373 (8.6) |  |
| Pakistani | 16,779 (5.6) | 126,346 (5.4) |  | 16,182 (5.6) | 16,182 (5.6) |  | 7,591 (6.0) | 126,346 (5.4) |  | 7,228 (6.0) | 7,228 (6.0) |  |
| Qatari | 36,177 (12.0) | 235,972 (10.1) |  | 36,091 (12.4) | 36,091 (12.4) |  | 18,940 (14.9) | 235,972 (10.1) |  | 18,813 (15.6) | 18,813 (15.6) |  |
| Sri Lankan | 9,598 (3.2) | 59,805 (2.6) |  | 9,249 (3.2) | 9,249 (3.2) |  | 3,190 (2.5) | 59,805 (2.6) |  | 2,950 (2.5) | 2,950 (2.5) |  |
| Sudanese | 8,231 (2.7) | 55,207 (2.4) |  | 8,017 (2.8) | 8,017 (2.8) |  | 3,638 (2.9) | 55,207 (2.4) |  | 3,430 (2.9) | 3,430 (2.9) |  |
| Other nationalities¶ | 48,201 (16.0) | 565,244 (24.3) |  | 43,703 (15.0) | 43,703 (15.0) |  | 25,719 (20.2) | 565,244 (24.3) |  | 22,692 (18.8) | 22,692 (18.8) |  |
| Comorbidity count |  |  |  |  |  |  |  |  |  |  |  |  |
| None | 241,571 (80.0) | 1,991,109 (85.5) | 0.15 | 235,153 (80.9) | 235,153 (80.9) | 0.00 | 104,077 (81.9) | 1,991,109 (85.5) | 0.15 | 100,533 (83.4) | 100,533 (83.4) | 0.00 |
| 1-2 | 48,776 (16.2) | 268,390 (11.5) |  | 45,342 (15.6) | 45,342 (15.6) |  | 20,547 (16.2) | 268,390 (11.5) |  | 18,271 (15.2) | 18,271 (15.2) |  |
| 3+ | 11,596 (3.8) | 69,540 (3.0) |  | 10,143 (3.5) | 10,143 (3.5) |  | 2,451 (1.9) | 69,540 (3.0) |  | 1,679 (1.4) | 1,679 (1.4) |  |

IQR denotes interquartile range and SMD standardized mean difference.
\*Individuals with a documented primary SARS-CoV-2 infection were exact-matched in a 1:1 ratio by sex, 10-year age group, nationality, comorbidity count, and calendar week of the SARS-CoV-2 test to the first eligible infection-naïve individual.
†SMD is the difference in the mean of a covariate between groups divided by the pooled standard deviation. An SMD <0.1 indicates adequate matching.
‡SMD is for the mean difference between groups divided by the pooled standard deviation.
§Nationalities were chosen to represent the most populous groups in Qatar.
¶These comprise 150 other nationalities in the unmatched primary-infection cohort and 183 other nationalities in the unmatched infection-naïve cohort, and 133 other nationalities in the matched primary-infection cohort and 133 other nationalities in the matched infection-naïve cohort in the Pre-Omicron Reinfection Study. These also comprise 145 other nationalities in the unmatched primary-infection cohort and 183 other nationalities in the unmatched infection-naïve cohort, and 123 other nationalities in the matched primary-infection cohort and 123 other nationalities in the matched infection-naïve cohort in the Omicron Reinfection Study.

There were 1,806 reinfections in the primary-infection cohort during follow-up, of which 6 progressed to severe and 1 to fatal COVID-19 (Figure S1). Meanwhile, there were 11,957 infections in the infection-naïve cohort, of which 297 progressed to severe, 19 to critical, and 12 to fatal COVID-19. Cumulative incidence of infection was 1.7% (95% CI: 1.6-1.8%) for the primary-infection cohort and 9.6% (95% CI: 9.4-9.9%) for the infection-naïve cohort, 15 months after the primary infection (Figure 1A).

**Figure 1.**
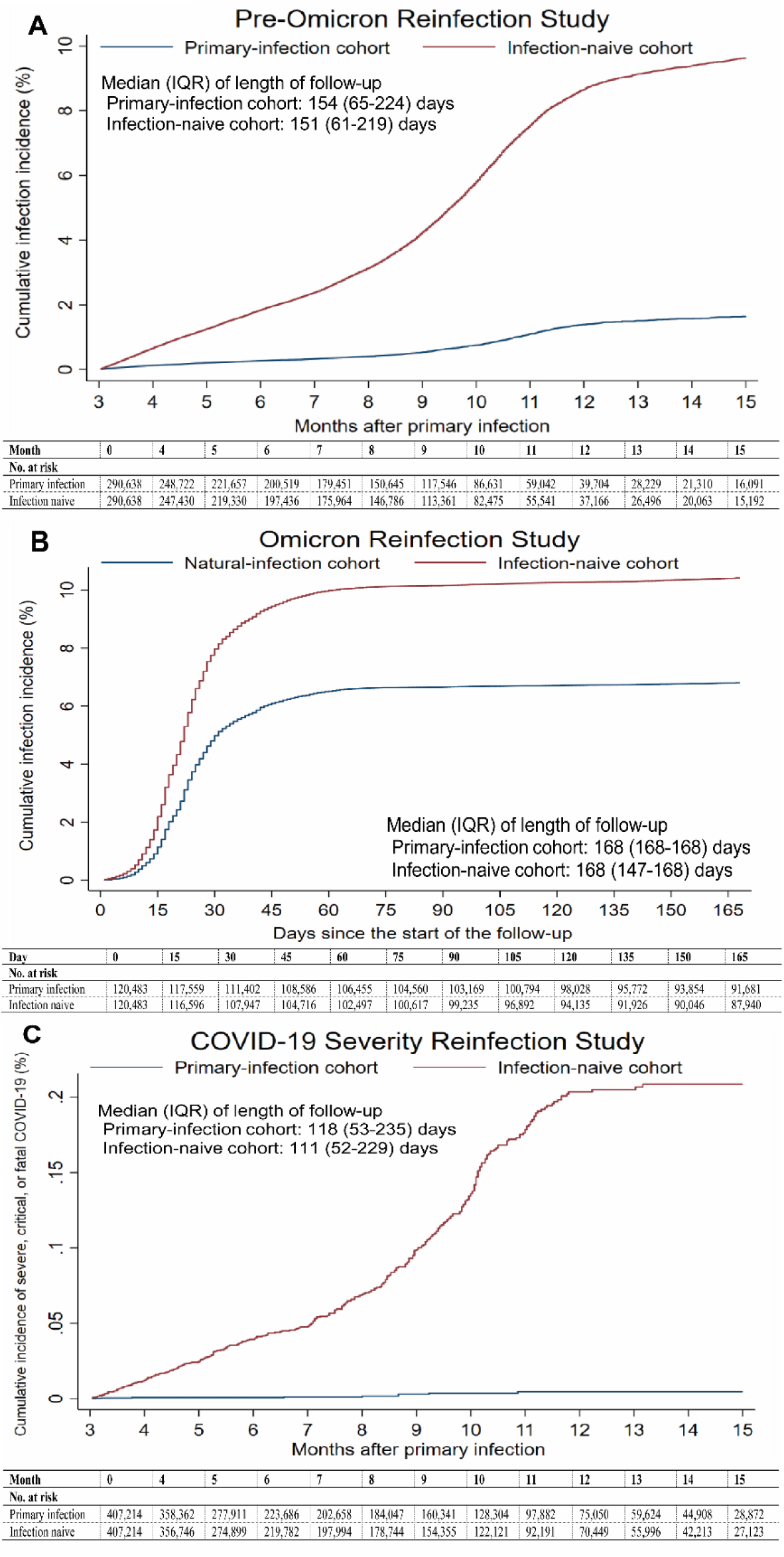
A) Cumulative incidence of infection in the Pre-Omicron Reinfection Study. B) Cumulative incidence of infection in the Omicron Reinfection Study. C) Cumulative incidence of severe, critical or fatal COVID-19 in the COVID-19 Severity Reinfection Study.

The overall hazard ratio for infection, adjusted for sex, 10-year age group, 10 nationality groups, comorbidity count, and SARS-CoV-2 test calendar week, was 0.14 (95% CI: 0.14-0.15; Table 3). Effectiveness of pre-Omicron primary infection against pre-Omicron reinfection was 85.5% (95% CI: 84.8-86.2%). Effectiveness increased slowly after the primary infection and reached 90.5% (95% CI: 88.4-92.3%) in the 7^th^ month after the primary infection (Figure 2A). Starting in the 8^th^ month, effectiveness waned slowly and reached ∼70% by the 16^th^ month. Fitting the waning of protection to a Gompertz curve suggested that effectiveness reaches 50% in the 22^nd^ month and <10% by the 32^nd^ month (Figure 3).

**Figure 2.**
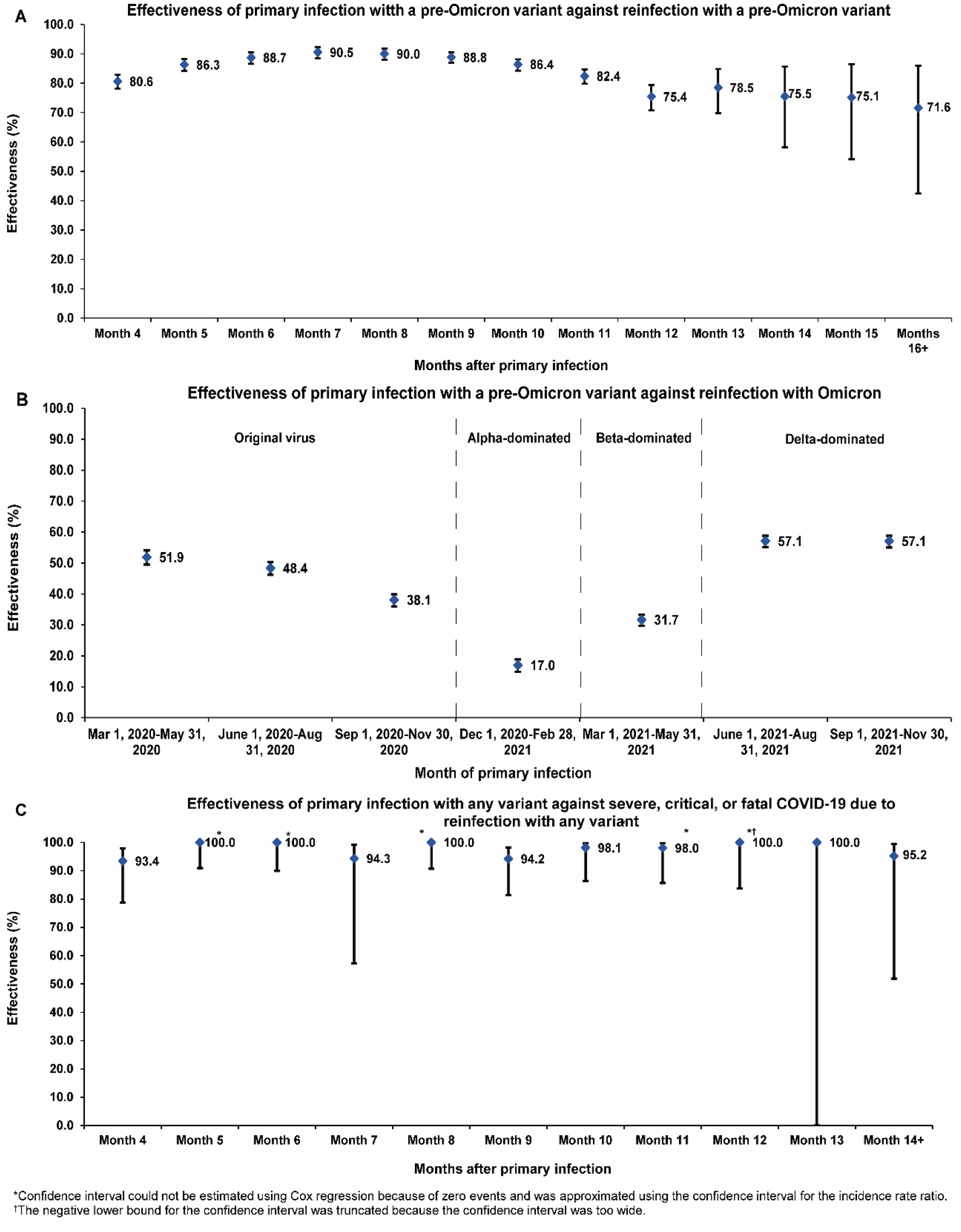
A) Effectiveness of pre-Omicron primary infection against pre-Omicron reinfection. B) Effectiveness of pre-Omicron primary infection against Omicron reinfection. C) Effectiveness of primary infection with any variant against severe, critical, or fatal COVID-19 due to reinfection with any variant.

**Figure 3.**
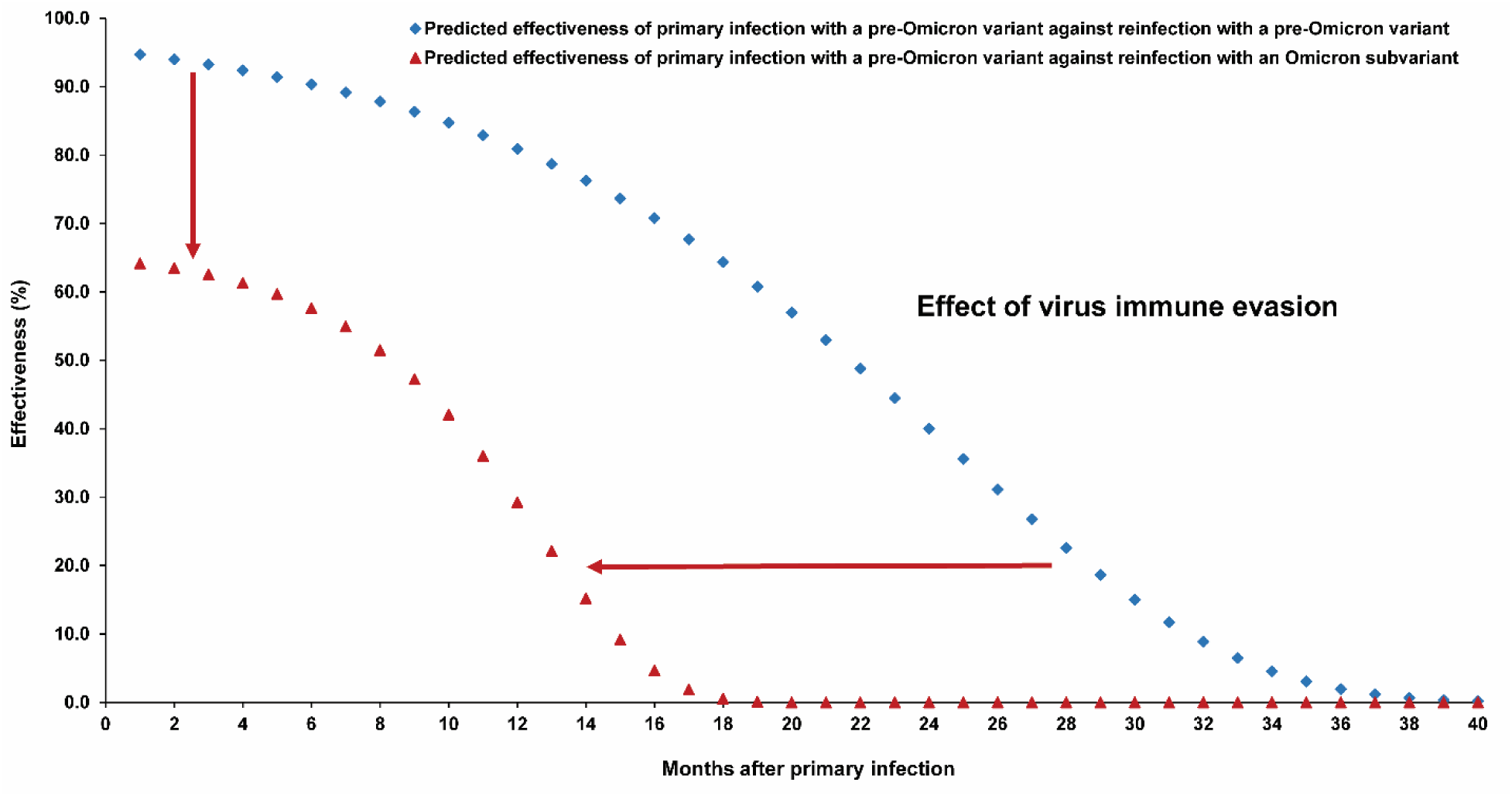
Extrapolated effectiveness of pre-Omicron primary infection against pre- Omicron reinfection, and extrapolated effectiveness of pre-Omicron primary infection against Omicron reinfection.

**Table 3.** Hazard ratios for the incidence of SARS-CoV-2 infection and incidence of severe, critical, or fatal COVID-19 in the Pre-Omicron Reinfection Study, Pre-Omicron Reinfection Study, and COVID-19 Severity Reinfection Study.

| Epidemiological measure | Primary-infection cohort | Infection-naïve cohort |
| --- | --- | --- |
| **Pre-Omicron Reinfection Study** |  |  |
| **Primary outcome** |  |  |
| Total follow-up time (person-weeks) | 6,578,466 | 6,432,430 |
| Incidence rate of infection (per 10,000 person-weeks) | 2.8 (2.6 to 2.9) | 18.6 (18.3 to 18.9) |
| Unadjusted hazard ratio for SARS-CoV-2 infection (95% CI) | 0.15 (0.14 to 0.15) |  |
| Adjusted hazard ratio for SARS-CoV-2 infection* (95% CI) | 0.14 (0.14 to 0.15) |  |
| Effectiveness against SARS-CoV-2 infection in % (95% CI) | 85.5 (84.8 to 86.2) |  |
| **Secondary outcome** |  |  |
| Unadjusted hazard ratio for severe, critical, or fatal COVID-19† (95% CI) | 0.02 (0.01 to 0.04) |  |
| Adjusted hazard ratio for severe, critical, or fatal COVID-19*† (95% CI) | 0.02 (0.01 to 0.04) |  |
| Effectiveness against severe, critical, or fatal COVID-19*† (95% CI) | 98.0 (95.7 to 99.0) |  |
| **Omicron Reinfection Study** |  |  |
| **Primary outcome** |  |  |
| Total follow-up time (person-weeks) | 2,493,870 | 2,411,571 |
| Incidence rate of infection (per 10,000 person-weeks) | 32.1 (31.4 to 32.8) | 50.7 (49.8 to 51.6) |
| Unadjusted hazard ratio for SARS-CoV-2 infection (95% CI) | 0.64 (0.62 to 0.66) |  |
| Adjusted hazard ratio for SARS-CoV-2 infection* (95% CI) | 0.62 (0.60 to 0.64) |  |
| Effectiveness against SARS-CoV-2 infection in % (95% CI) | 38.1 (36.3 to 39.8) |  |
| **Secondary outcome** |  |  |
| Unadjusted hazard ratio for severe, critical, or fatal COVID-19† (95% CI) | 0.13 (0.05 to 0.33) |  |
| Adjusted hazard ratio for severe, critical, or fatal COVID-19*† (95% CI) | 0.11 (0.04 to 0.29) |  |
| Effectiveness against severe, critical, or fatal COVID-19*† (95% CI) | 88.6 (70.9 to 95.5) |  |
| **Reinfection COVID-19 Severity Study** |  |  |
| **Primary outcome** |  |  |
| Total follow-up time (person-weeks) | 9,290,507 | 9,022,235 |
| Incidence rate of severe, critical, or fatal COVID-19 (per 10,000 person-weeks) | 0.01 (0.01 to 0.02) | 0.40 (0.36 to 0.44) |
| Unadjusted hazard ratio for severe, critical, or fatal COVID-19† (95% CI) | 0.03 (0.01 to 0.05) |  |
| Adjusted hazard ratio for severe, critical, or fatal COVID-19*† (95% CI) | 0.03 (0.01 to 0.05) |  |
| Effectiveness against severe, critical, or fatal COVID-19*† (95% CI) | 97.3 (94.9 to 98.6) |  |
| **Secondary outcome** |  |  |
| Unadjusted hazard ratio for SARS-CoV-2 infection (95% CI) | 0.31 (0.30 to 0.32) |  |
| Adjusted hazard ratio for SARS-CoV-2 infection* (95% CI) | 0.31 (0.30 to 0.31) |  |
| Effectiveness against SARS-CoV-2 infection in % (95% CI) | 69.4 (68.6 to 70.3) |  |

CI denotes confidence interval, COVID-19 coronavirus disease 2019, and SARS-CoV-2 severe acute respiratory syndrome coronavirus 2.
\*Cox regression analysis adjusted for sex, 10-year age group (Table 1), 10 nationality groups (Table 1), comorbidity count (Table 1), and calendar week of the SARS-CoV-2 test.
†Severity,<sup>22</sup> criticality,<sup>22</sup> and fatality<sup>23</sup> were defined according to the World Health Organization guidelines.

Effectiveness of pre-Omicron primary infection against severe, critical, or fatal COVID-19 due to pre-Omicron reinfection was 98.0% (95% CI: 95.7-99.0%; Table 3). In the additional analysis restricting the matched cohorts to the sub-cohorts ≥50 years of age (25,595 individuals), effectiveness against reinfection and against severe, critical, or fatal COVID-19 reinfection was 90.7% (95% CI: 88.4-92.5%) and 97.4% (95% CI: 91.9-99.2%), respectively. In the sensitivity analysis adjusting the overall hazard ratio by the ratio of testing frequency, effectiveness against reinfection was 79.5% (95% CI: 78.4-80.5%). More results are in Section S3.

### Omicron Reinfection Study

Figure S2 shows the process of population selection and Table 1 describes the baseline characteristics of the full and matched cohorts. The matched cohorts each included 120,483 individuals. The cohorts are representative of Qatar’s population (Table S2).

There were 7,995 reinfections in the primary-infection cohort during follow-up, of which 5 progressed to severe COVID-19 (Figure S2). Meanwhile, there were 12,230 infections in the infection-naïve cohort, of which 26 progressed to severe, 7 to critical, and 5 to fatal COVID-19. Cumulative incidence of infection was 6.8% (95% CI: 6.7-6.9%) for the primary-infection cohort and 10.4% (95% CI: 10.2-10.6%) for the infection-naïve cohort, 165 days after the start of follow-up (Figure 1B).

The overall adjusted hazard ratio for infection was 0.62 (95% CI: 0.60-0.64; Table 3). Effectiveness of pre-Omicron primary infection against Omicron reinfection was 38.1% (95% CI: 36.3-39.8%). Effectiveness varied for the primary-infection sub-cohorts (Figure 2B). It was ∼60% for those with a more recent primary infection, between June 1, 2021 and November 30, 2021, during Delta-dominated incidence.^4, 34, 35^ Effectiveness declined with time since primary infection and was 17.0% (95% CI: 10.1-23.5%) for those with a primary infection between December 1, 2020 and February 28, 2021, during Alpha-dominated incidence.^4, 34, 35^ However, higher effectiveness of ∼50% was estimated for those with a primary infection before August 31, 2020, during original-virus incidence (note discussion in Section S3).^4, 34, 35^ Fitting the waning of protection to a Gompertz curve suggested that effectiveness reaches 50% in the 8^th^ month after primary infection and <10% by the 15^th^ month (Figure 3).

Effectiveness of pre-Omicron primary infection against severe, critical, or fatal COVID-19 due to Omicron reinfection was 88.6% (95% CI: 70.9-95.5%; Table 3). In the additional analysis restricting the matched cohorts to the sub-cohorts ≥50 years of age (6,304 individuals), effectiveness against reinfection and against severe, critical, or fatal COVID-19 reinfection was 21.6% (95% CI: 11.1-31.0%) and 84.6% (95% CI: 59.7-94.1%), respectively. In the sensitivity analysis adjusting the overall hazard ratio by the ratio of testing frequency, effectiveness against reinfection was 31.7% (95% CI: 29.7-33.6%). More results are in Section S3.

### COVID-19 Severity Reinfection Study

Figure S3 shows the process of population selection and Table 2 describes baseline characteristics of the full and matched cohorts. The matched cohorts each included 407,214 individuals, representative of Qatar’s population (Table S2).

**Table 2.** Baseline characteristics of the eligible and matched primary-infection and infection-naive cohorts in the COVID-19 Severity Reinfection Study.

| Characteristics | Full eligible cohorts |  |  | Matched cohorts* |  |  |
| --- | --- | --- | --- | --- | --- | --- |
|  | Primary-infection cohort | Infection-naïve cohort | SMD† | Primary-infection cohort | Infection-naïve cohort | SMD† |
|  | N=438,854 | N=2,843,496 |  | N=407,214 | N=407,214 |  |
| Median age (IQR)—years | 30 (16-39) | 31 (23-39) | 0.15‡ | 30 (18-38) | 30 (19-38) | 0.00‡ |
| Age group |  |  |  |  |  |  |
| 0-9 years | 75,287 (17.2) | 324,956 (11.4) |  | 69,059 (17.0) | 69,059 (17.0) |  |
| 10-19 years | 42,913 (9.8) | 196,544 (6.9) |  | 36,029 (8.9) | 36,029 (8.9) |  |
| 20-29 years | 97,277 (22.2) | 748,686 (26.3) |  | 92,980 (22.8) | 92,980 (22.8) |  |
| 30-39 years | 122,856 (28.0) | 880,600 (31.0) | 0.21 | 117,269 (28.8) | 117,269 (28.8) | 0.00 |
| 40-49 years | 64,930 (14.8) | 429,083 (15.1) |  | 60,769 (14.9) | 60,769 (14.9) |  |
| 50-59 years | 25,272 (5.8) | 179,111 (6.3) |  | 22,858 (5.6) | 22,858 (5.6) |  |
| 60-69 years | 7,811 (1.8) | 64,210 (2.3) |  | 6,492 (1.6) | 6,492 (1.6) |  |
| 70+ years | 2,508 (0.6) | 20,306 (0.7) |  | 1,758 (0.4) | 1,758 (0.4) |  |
| Sex |  |  |  |  |  |  |
| Male | 297,317 (67.8) | 1,979,916 (69.6) | 0.04 | 280,419 (68.9) | 280,419 (68.9) | 0.00 |
| Female | 141,537 (32.2) | 863,580 (30.4) |  | 126,795 (31.1) | 126,795 (31.1) |  |
| Nationality§ |  |  |  |  |  |  |
| Bangladeshi | 29,599 (6.7) | 175,765 (6.2) |  | 27,691 (6.8) | 27,691 (6.8) |  |
| Egyptian | 23,003 (5.2) | 136,182 (4.8) |  | 21,534 (5.3) | 21,534 (5.3) |  |
| Filipino | 32,871 (7.5) | 200,002 (7.0) |  | 31,256 (7.7) | 31,256 (7.7) |  |
| Indian | 108,178 (24.7) | 804,623 (28.3) |  | 107,720 (26.5) | 107,720 (26.5) |  |
| Nepalese | 43,108 (9.8) | 253,949 (8.9) | 0.26 | 40,098 (9.9) | 40,098 (9.9) | 0.00 |
| Pakistani | 22,647 (5.2) | 170,868 (6.0) |  | 21,966 (5.4) | 21,966 (5.4) |  |
| Qatari | 71,132 (16.2) | 249,740 (8.8) |  | 60,438 (14.8) | 60,438 (14.8) |  |
| Sri Lankan | 11,912 (2.7) | 79,189 (2.8) |  | 11,519 (2.8) | 11,519 (2.8) |  |
| Sudanese | 11,666 (2.7) | 62,039 (2.2) |  | 10,914 (2.7) | 10,914 (2.7) |  |
| Other nationalities¶ | 84,738 (19.3) | 711,139 (25.0) |  | 74,078 (18.2) | 74,078 (18.2) |  |
| Comorbidity count |  |  |  |  |  |  |
| None | 352,738 (80.4) | 2,496,024 (87.8) |  | 334,409 (82.1) | 334,409 (82.1) |  |
| 1-2 | 72,153 (16.4) | 277,344 (9.8) | 0.21 | 61,720 (15.2) | 61,720 (15.2) | 0.00 |
| 3+ | 13,963 (3.2) | 70,128 (2.5) |  | 11,085 (2.7) | 11,085 (2.7) |  |

IQR denotes interquartile range and SMD standardized mean difference.
†Individuals with a documented primary SARS-CoV-2 infection were exact-matched in a 1:1 ratio by sex, 10-year age group, nationality, comorbidity count, and calendar week of the SARS-CoV-2 test to the first eligible infection-naïve individual.
‡SMD is the difference in the mean of a covariate between groups divided by the pooled standard deviation. An SMD <0.1 indicates adequate matching.
§SMD is for the mean difference between groups divided by the pooled standard deviation.
¶Nationalities were chosen to represent the most populous groups in Qatar.
\*These comprise 161 other nationalities in the unmatched primary-infection cohort and 183 other nationalities in the unmatched infection-naïve cohort, and 141 other nationalities in the matched primary-infection cohort and 141 other nationalities in the matched infection-naïve cohort.

There were 7,082 reinfections in the primary-infection cohort during follow-up, of which 9 progressed to severe and 1 progressed to fatal COVID-19 (Figure S3). Meanwhile, there were 21,645 infections in the infection-naïve cohort, of which 315 progressed to severe, 25 to critical, and 18 to fatal COVID-19. Cumulative incidence of severe, critical, or fatal COVID-19 was 0.00% (95% CI: 0.00-0.01%) for the primary-infection cohort and 0.21% (95% CI: 0.19-0.23%) for the infection-naïve cohort, 15 months after the primary infection (Figure 1C).

The overall adjusted hazard ratio for severe, critical, or fatal COVID-19 was estimated at 0.03 (95% CI: 0.01-0.05; Table 3). Effectiveness of primary infection with any variant against severe, critical, or fatal COVID-19 due to reinfection with any variant was 97.3% (95% CI: 94.9- 98.6%). Variation by month after primary infection was negligible, with no evidence for waning (Figure 3C). Effectiveness was ∼100% up to the 14^th^ month after primary infection.

Effectiveness of primary infection with any variant against reinfection with any variant was 69.4% (95% CI: 68.6-70.3%; Table 3). In the additional analysis restricting the matched cohorts to the sub-cohorts ≥50 years of age (31,108 individuals), effectiveness against reinfection and against severe, critical, or fatal COVID-19 was 75.3% (95% CI: 72.0-78.2%) and 95.4% (95% CI: 89.4-98.0%), respectively. More results are in Section S3.

## Discussion

Protection of natural infection waned with time after primary infection, prior to Omicron emergence, and reached ∼70% by the 16^th^ month. This waning likely reflects genuine waning in biological immunity rather than viral immune evasion, as pre-Omicron variants demonstrated much less immune evasion than Omicron.^14–16^ This waning in natural immunity mirrors that of vaccine immunity,^4, 6, 30^ but at a slower rate. Vaccine immunity may last for only a year,^4, 6, 30^ but natural immunity, assuming Gompertz decay, may last for 3 years, as also suggested by long-term follow-up of SARS-CoV-1-associated antibodies,^36^ and incidentally not dissimilar to pandemic-influenza-associated antibodies.^37^

Immune evasion of Omicron subvariants reduced overall protection of pre-Omicron natural immunity and accelerated its waning (Figure 3), mirroring the effect of Omicron on vaccine immunity, but at a slower rate. Vaccine immunity against Omicron subvariants lasts for <6 months,^5, 7, 8^ but pre-Omicron natural immunity, assuming Gompertz decay, may last for just over a year.

Despite waning protection against reinfection, strikingly, there was no evidence for waning of protection against severe COVID-19 at reinfection. This remained ∼100%, even 14 months after the primary infection, with no appreciable effect for Omicron immune evasion in reducing it.

This pattern also mirrors that of vaccine immunity, which wanes rapidly against infection, but is durable against severe COVID-19, regardless of variant.^4, 6–8, 30^

Infection with common-cold coronaviruses, and perhaps influenza,^38^ induces only a year-long immunity against reinfection,^10^ but life-long immunity against severe reinfection.^2^ To what extent this pattern reflects waning in biological immunity or immune evasion with virus evolution over the global season is unclear. Above results suggest that SARS-CoV-2 may exhibit a similar pattern to that of common-cold coronaviruses within few years. Short-term biological immunity against reinfection of 3 years may decline as a result of viral evolution and immune evasion, leading to periodic (possibly annual) waves of infection. However, the lasting immunity against severe reinfection will contribute to a pattern of benign infection. Most primary infections would occur in childhood and would likely not be severe. Adults would only experience periodic reinfections, also not likely to be severe.

This study has limitations. We investigated incidence of documented infections, but other infections may have occurred and gone undocumented. Undocumented infections confer immunity or boost existing immunity, thereby perhaps affecting the estimates (note Section S3).

Differences in testing frequency existed between the followed cohorts, but these were small and adjusted estimates in sensitivity analyses confirmed similar findings. Gompertz function^33^ was used to parametrize immunity decay, based on empiric goodness-of-fit, but this analysis serves only as an informed exploratory extrapolation that remains to be confirmed with more follow-up of cohorts. With Qatar’s young population, our findings may not be generalizable to other countries where elderly citizens constitute a larger proportion of the total population. However, additional analyses restricting the matched cohorts to those ≥50 years of age showed findings resembling those for the total population.

Depletion of the primary-infection cohorts by COVID-19 mortality at time of primary infection may have biased these cohorts toward healthier individuals with stronger immune responses.

However, COVID-19 mortality has been low in Qatar’s predominantly young population,^19, 39^ totaling 679 COVID-19 deaths (<0.1% of primary infections) up to June 29, 2022, and much smaller than the size of the study cohorts. A survival effect seems unlikely to explain or appreciably affect study findings, apart perhaps from protection against severe COVID-19.

As an observational study, investigated cohorts were neither blinded nor randomized, so unmeasured or uncontrolled confounding cannot be excluded. While matching was done for sex, age, nationality, comorbidity count, and timing of primary infection, this was not possible for other factors such as geography or occupation, as such data were unavailable. However, Qatar is essentially a city state and infection incidence was broadly distributed across neighborhoods.

Nearly 90% of Qatar’s population are expatriates from over 150 countries, coming here because of employment.^19^ Most are craft and manual workers working in development projects.^19^ Nationality, age, and sex provide a powerful proxy for socio-economic status in this country.^19, 24–27^ Nationality alone is strongly associated with occupation.^19, 25–27^

Matching was done to control for factors that affect infection exposure in Qatar.^19, 24–27^ The matching prescription had already been investigated in previous studies of different epidemiologic designs, and using control groups to test for null effects.^4, 28–31^ These control groups included unvaccinated cohorts versus vaccinated cohorts within two weeks of the first dose,^4, 28–30^ when vaccine protection is negligible,^40^ and mRNA-1273- versus BNT162b2- vaccinated cohorts, also in the first two weeks after the first dose.^31^ These studies have shown that this prescription provides adequate control of the differences in infection exposure.^4, 28–31^ These analyses were implemented using Qatar’s total population with large sample sizes, thus minimizing the likelihood of bias.

In conclusion, protection of natural infection against reinfection wanes and may diminish within a few years. Omicron immune evasion accelerates this waning. Meanwhile, protection against severe reinfection is very strong with no evidence for waning, regardless of variant, for over 14 months after the primary infection.

## Data Availability

The dataset of this study is a property of the Qatar Ministry of Public Health that was provided to the researchers through a restricted-access agreement that prevents sharing the dataset with a third party or publicly. Future access to this dataset can be considered through a direct application for data access to Her Excellency the Minister of Public Health (https://www.moph.gov.qa/english/Pages/default.aspx). Aggregate data are available within the manuscript and its Supplementary information.

## Sources of support and acknowledgements

We acknowledge the many dedicated individuals at Hamad Medical Corporation, the Ministry of Public Health, the Primary Health Care Corporation, Qatar Biobank, Sidra Medicine, and Weill Cornell Medicine-Qatar for their diligent efforts and contributions to make this study possible.

The authors are grateful for institutional salary support from the Biomedical Research Program and the Biostatistics, Epidemiology, and Biomathematics Research Core, both at Weill Cornell Medicine-Qatar, as well as for institutional salary support provided by the Ministry of Public Health, Hamad Medical Corporation, and Sidra Medicine. The authors are also grateful for the Qatar Genome Programme and Qatar University Biomedical Research Center for institutional support for the reagents needed for the viral genome sequencing. The funders of the study had no role in study design, data collection, data analysis, data interpretation, or writing of the article.

Statements made herein are solely the responsibility of the authors.

## Author contributions

HC co-designed the study, performed the statistical analyses, and co-wrote the first draft of the article. LJA conceived and co-designed the study, led the statistical analyses, and co-wrote the first draft of the article. NG provided technical advice and insights about immunity decay. PT and MRH conducted multiplex, RT-qPCR variant screening and viral genome sequencing. HY, HAK, and MS conducted viral genome sequencing. All authors contributed to data collection and acquisition, database development, discussion and interpretation of the results, and to the writing of the manuscript. All authors have read and approved the final manuscript.

## Section S1. Laboratory methods and variant ascertainment

### Real-time reverse-transcription polymerase chain reaction testing

Nasopharyngeal and/or oropharyngeal swabs were collected for polymerase chain reaction (PCR) testing and placed in Universal Transport Medium (UTM). Aliquots of UTM were: 1) extracted on KingFisher Flex (Thermo Fisher Scientific, USA), MGISP-960 (MGI, China), or ExiPrep 96 Lite (Bioneer, South Korea) followed by testing with real-time reverse-transcription PCR (RT-qPCR) using TaqPath COVID-19 Combo Kits (Thermo Fisher Scientific, USA) on an ABI 7500 FAST (Thermo Fisher Scientific, USA); 2) tested directly on the Cepheid GeneXpert system using the Xpert Xpress SARS-CoV-2 (Cepheid, USA); or 3) loaded directly into a Roche cobas 6800 system and assayed with the cobas SARS-CoV-2 Test (Roche, Switzerland). The first assay targets the viral S, N, and ORF1ab gene regions. The second targets the viral N and E- gene regions, and the third targets the ORF1ab and E-gene regions.

All PCR testing was conducted at the Hamad Medical Corporation Central Laboratory or Sidra Medicine Laboratory, following standardized protocols.

### Rapid antigen testing

Severe acute respiratory syndrome coronavirus 2 (SARS-CoV-2) antigen tests were performed on nasopharyngeal swabs using one of the following lateral flow antigen tests: Panbio COVID- 19 Ag Rapid Test Device (Abbott, USA); SARS-CoV-2 Rapid Antigen Test (Roche, Switzerland); Standard Q COVID-19 Antigen Test (SD Biosensor, Korea); or CareStart COVID- 19 Antigen Test (Access Bio, USA). All antigen tests were performed point-of-care according to each manufacturer’s instructions at public or private hospitals and clinics throughout Qatar with prior authorization and training by the Ministry of Public Health (MOPH). Antigen test results were electronically reported to the MOPH in real time using the Antigen Test Management System which is integrated with the national Coronavirus Disease 2019 (COVID-19) database.

### Classification of infections by variant type

Surveillance for SARS-CoV-2 variants in Qatar is based on viral genome sequencing and multiplex RT-qPCR variant screening^1^ of random positive clinical samples,^2–7^ complemented by deep sequencing of wastewater samples.^4, 8^ Further details on the viral genome sequencing and multiplex RT-qPCR variant screening throughout the SARS-CoV-2 waves in Qatar can be found in previous publications.^2–7, 9–14^

## Section S2. COVID-19 severity, criticality, and fatality classification

Classification of COVID-19 case severity (acute-care hospitalizations),^15^ criticality (intensive-care-unit hospitalizations),^15^ and fatality^16^ followed World Health Organization (WHO) guidelines. Assessments were made by trained medical personnel independent of study investigators and using individual chart reviews, as part of a national protocol applied to every hospitalized COVID-19 patient. Each hospitalized COVID-19 patient underwent an infection severity assessment every three days until discharge or death. We classified individuals who progressed to severe, critical, or fatal COVID-19 between the time of the documented infection and the end of the study based on their worst outcome, starting with death,^16^ followed by critical disease,^15^ and then severe disease.^15^

Severe COVID-19 disease was defined per WHO classification as a SARS-CoV-2 infected person with “oxygen saturation of <90% on room air, and/or respiratory rate of >30 breaths/minute in adults and children >5 years old (or ≥60 breaths/minute in children <2 months old or ≥50 breaths/minute in children 2-11 months old or ≥40 breaths/minute in children 1–5 years old), and/or signs of severe respiratory distress (accessory muscle use and inability to complete full sentences, and, in children, very severe chest wall indrawing, grunting, central cyanosis, or presence of any other general danger signs)”.^15^ Detailed WHO criteria for classifying SARS-CoV-2 infection severity can be found in the WHO technical report.^15^

Critical COVID-19 disease was defined per WHO classification as a SARS-CoV-2 infected person with “acute respiratory distress syndrome, sepsis, septic shock, or other conditions that would normally require the provision of life sustaining therapies such as mechanical ventilation (invasive or non-invasive) or vasopressor therapy”.^15^ Detailed WHO criteria for classifying SARS-CoV-2 infection criticality can be found in the WHO technical report.^15^

COVID-19 death was defined per WHO classification as “a death resulting from a clinically compatible illness, in a probable or confirmed COVID-19 case, unless there is a clear alternative cause of death that cannot be related to COVID-19 disease (e.g. trauma). There should be no period of complete recovery from COVID-19 between illness and death. A death due to COVID- 19 may not be attributed to another disease (e.g. cancer) and should be counted independently of preexisting conditions that are suspected of triggering a severe course of COVID-19”. Detailed WHO criteria for classifying COVID-19 death can be found in the WHO technical report.^16^

## Section S3. Additional material for the Results section Pre-Omicron Reinfection Study

The median time of follow-up was 154 days (interquartile range (IQR), 65-224 days) for the primary-infection cohort and 151 days (IQR, 61-219 days) for the infection-naïve cohort (Figure 1A). The proportion of individuals who had a SARS-CoV-2 test during follow-up was 29.7% for the primary-infection cohort and 36.4% for the infection-naïve cohort. The testing frequency was 0.56 and 0.79 tests per person, respectively.

The pattern of waning of protection in Figure 2A was fitted to a Gompertz function,^17^ but after setting the effectiveness values in months 4-6 after primary infection at 90.5%, the value at the 7^th^ month. This was done to correct for the likely underestimation of the observed effectiveness in these months because of the effect of PCR-positive tests that reflected prolonged infections as opposed to true reinfections.^18–23^ The fitted Gompertz function suggested that effectiveness against reinfection reaches 50% in the 22^nd^ month after primary infection, and reaches <10% by the 32^nd^ month (Figure 3).

### Omicron Reinfection Study

The median time of follow-up was 168 days (IQR, 168-168 days) for the primary-infection cohort and 168 days (IQR, 147-168 days) for the infection-naïve cohort (Figure 1B). The proportion of individuals who had a SARS-CoV-2 test during follow-up was 29.9% for the primary-infection cohort and 32.5% for the infection-naïve cohort. The testing frequency was 0.50 and 0.55 tests per person, respectively.

The pattern of waning of protection in Figure 2B was fitted to a Gompertz function,^17^ but after excluding the effectiveness values before December 1, 2020. Incidence before this date, that is during the first wave, affected mainly the craft and manual workers population of Qatar, that constitutes 60% of the total population and where SARS-CoV-2 seroprevalence exceeded 50% by the end of the first wave.^24–28^ This population segment has also the lowest testing rates in the population.^29^ It is possible that many of these workers may have had mild or asymptomatic reinfections^30, 31^ that were not documented during the four subsequent waves.^11, 12, 32, 33^ This undocumented immune boosting, subsequent to the primary infection, may explain the higher-than-expected effectiveness for those who had their primary infection prior to December 1, 2020. The fitted Gompertz function suggested that effectiveness against reinfection reaches 50% in the 8^th^ month after primary infection, and reaches <10% by the 15^th^ month (Figure 3).

### COVID-19 Severity Reinfection Study

The median time of follow-up was 118 days (IQR, 53-235 days) for the primary-infection cohort and 111 days (IQR, 52-229 days) for the infection-naïve cohort (Figure 1C). The proportion of individuals who had a SARS-CoV-2 test during follow-up was 28.0% for the primary-infection cohort and 31.7% for the infection-naïve cohort. The testing frequency was 0.56 and 0.70 tests per person, respectively.

Since there was no evidence for waning in effectiveness against severe, critical, or fatal COVID- 19 due to reinfection, there was no relevance to fit a Gompertz function to the effectiveness trend after primary infection.

---

**Table S1.** STROBE checklist for cohort studies.

|  | Item No | Recommendation | Main Text page |
| --- | --- | --- | --- |
| Title and abstract | 1 | (a) Indicate the study’s design with a commonly used term in the title or the abstract<br>(b) Provide in the abstract an informative and balanced summary of what was done and what was found | Abstract<br>Abstract |
| Introduction |  |  |  |
| Background/rationale | 2 | Explain the scientific background and rationale for the investigation being reported | Introduction |
| Objectives | 3 | State specific objectives, including any prespecified hypotheses | Introduction |
| Methods |  |  |  |
| Study design | 4 | Present key elements of study design early in the paper | Methods (‘Study designs and cohorts’) |
| Setting | 5 | Describe the setting, locations, and relevant dates, including periods of recruitment, exposure, follow-up, and data collection | Methods (‘Study designs and cohorts’, ‘Cohort matching and follow-up’, ‘Pre-Omicron Reinfection Study’, ‘Omicron Reinfection Study’, & ‘COVID-19 Severity Reinfection Study’) & Figures S1-S3 in Supplementary Appendix |
| Participants | 6 | (a) Give the eligibility criteria, and the sources and methods of selection of participants. Describe methods of follow-up<br>(b) For matched studies, give matching criteria and number of exposed and unexposed | Methods (‘Study designs and cohorts’, ‘Cohort matching and follow-up’, ‘Pre-Omicron Reinfection Study’, ‘Omicron Reinfection Study’, & ‘COVID-19 Severity Reinfection Study’) & Figures S1-S3 in Supplementary Appendix |
| Variables | 7 | Clearly define all outcomes, exposures, predictors, potential confounders, and effect modifiers. Give diagnostic criteria, if applicable | Methods (‘Study designs and cohorts’, ‘Cohort matching and follow-up’, ‘Pre-Omicron Reinfection Study’, ‘Omicron Reinfection Study’, & ‘COVID-19 Severity Reinfection Study’), Tables 1-2, & Sections S1 & S2 in Supplementary Appendix |
| Data sources/measurement | 8* | For each variable of interest, give sources of data and details of methods of assessment (measurement). Describe comparability of assessment methods if there is more than one group | Methods (‘Study population and data sources’ & ‘Statistical analysis’), Tables 1-2, & Sections S1 & S2 in Supplementary Appendix |
| Bias | 9 | Describe any efforts to address potential sources of bias | Methods (‘Cohort matching and follow-up’) |
| Study size | 10 | Explain how the study size was arrived at | Figures S1-S3 in Supplementary Appendix |
| Quantitative variables | 11 | Explain how quantitative variables were handled in the analyses. If applicable, describe which groupings were chosen and why | Methods (‘Cohort matching and follow-up’) & Tables 1-2 |
| Statistical methods | 12 | (a) Describe all statistical methods, including those used to control for confounding | Methods (‘Statistical analysis’) |
|  |  | (b) Describe any methods used to examine subgroups and interactions | Methods (‘Statistical analysis’, paragraph 3) |
|  |  | (c) Explain how missing data were addressed | NA, see Methods (‘Study population and data sources’) |
|  |  | (d) If applicable, explain how loss to follow-up was addressed | NA, see Methods (‘Study designs and cohorts’, paragraph 1) |
|  |  | (e) Describe any sensitivity analyses | Methods (‘Statistical analysis’, paragraph 3) |
| Results |  |  |  |
| Participants | 13* | (a) Report numbers of individuals at each stage of study—eg numbers potentially eligible, examined for eligibility, confirmed eligible, included in the study, completing follow-up, and analysed<br>(b) Give reasons for non-participation at each stage<br>(c) Consider use of a flow diagram | Figures S1-S3 in Supplementary Appendix |
| Descriptive data | 14 | (a) Give characteristics of study participants (eg demographic, clinical, social) and information on exposures and potential confounders | Tables 1-2 |
|  |  | (b) Indicate number of participants with missing data for each variable of interest | NA, see Methods (‘Study population and data sources’) |
|  |  | (c) Summarise follow-up time (eg, average and total amount) | Results ('Pre-Omicron Reinfection Study', 'Omicron Reinfection Study', & 'COVID-19 Severity Reinfection Study'), Figure 1, & Table 3 |
| Outcome data | 15 | Report numbers of outcome events or summary measures over time | Results ('Pre-Omicron Reinfection Study', 'Omicron Reinfection Study', & 'COVID-19 Severity Reinfection Study'), Figure 1, & Table 3 |
| Main results | 16 | (a) Give unadjusted estimates and, if applicable, confounder-adjusted estimates and their precision (eg, 95% confidence interval). Make clear which confounders were adjusted for and why they were included | Results ('Pre-Omicron Reinfection Study', 'Omicron Reinfection Study', & 'COVID-19 Severity Reinfection Study'), & Table 3 |
|  |  | (b) Report category boundaries when continuous variables were categorized | Tables 1-2 |
|  |  | (c) If relevant, consider translating estimates of relative risk into absolute risk for a meaningful time period | NA |
| Other analyses | 17 | Report other analyses done—eg analyses of subgroups and interactions, and sensitivity analyses | Results ('Pre-Omicron Reinfection Study', 'Omicron Reinfection Study', & 'COVID-19 Severity Reinfection Study') & Figures 2-3 |
| Discussion |  |  |  |
| Key results | 18 | Summarise key results with reference to study objectives | Discussion, paragraphs 1-4 |
| Limitations | 19 | Discuss limitations of the study, taking into account sources of potential bias or imprecision. Discuss both direction and magnitude of any potential bias | Discussion, paragraphs 5-8 |
| Interpretation | 20 | Give a cautious overall interpretation of results considering objectives, limitations, multiplicity of analyses, results from similar studies, and other relevant evidence | Discussion, paragraph 9 |
| Generalisability | 21 | Discuss the generalisability (external validity) of the study results | Discussion, paragraphs 5-7 and Table S2 in Supplementary Appendix |
| Other information |  |  |  |
| Funding | 22 | Give the source of funding and the role of the funders for the present study and, if applicable, for the original study on which the present article is based | Sources of support & acknowledgements |

**Table S2.** Representativeness of study participants.

|  |  |
| --- | --- |
| **Category** |  |
| Disease, problem, or condition under investigation | Protection conferred by natural SARS-CoV-2 infection against SARS-CoV-2 reinfection and against severe, critical, or fatal COVID-19 due to reinfection. |
| Special considerations related to |  |
| Sex and gender | Three national, matched, retrospective, target-trial cohort studies were conducted to compare incidence of SARS-CoV-2 infection and COVID-19 severity among those unvaccinated but with a documented SARS-CoV-2 primary infection, to incidence among those infection-naïve and unvaccinated. Cohorts were exact-matched by sex to control for potential differences in the risk of exposure to SARS-CoV-2 infection by sex. |
| Age | Cohorts were exact-matched by 10-year age group to control for potential differences in the risk of exposure to SARS-CoV-2 infection by age. Nonetheless, with the young population of Qatar, our findings may not be generalizable to other countries where elderly citizens constitute a larger proportion of the total population. Having said so, the additional analyses restricting the matched cohorts to those $\geq 50$ years of age showed similar findings to those for the total population. |
| Race or ethnicity group | Cohorts were exact-matched by nationality to control for potential differences in the risk of exposure to SARS-CoV-2 infection by nationality. Nationality is associated with race and ethnicity in the population of Qatar. |
| Geography | Individual-level data on geography were not available, but Qatar is essentially a city state and infection incidence was broadly distributed across the country's neighborhoods/areas. Cohorts were exact-matched by nationality to control for potential differences in the risk of exposure to SARS-CoV-2 infection by nationality. Qatar has unusually diverse demographics in that 89% of the population are international expatriate residents coming from over 150 countries from all world regions. |
| Other considerations | Individual-level data on occupation were not available but matching by nationality may have (partially) controlled the differences in occupational risk, in consideration of the association between nationality and occupation in Qatar. To ensure that all individuals in all cohorts have a record of active residence in Qatar at the same calendar time, individuals who were diagnosed with a primary infection in a specific week were matched to infection-naïve individuals who had a record of a SARS-CoV-2 negative test in that same calendar week. |
| Overall representativeness of this study | The study was based on the total population of Qatar and thus the study population is broadly representative of the diverse, by national background, but young and predominantly male, total population of Qatar. While there could be differences in the risk of exposure to SARS-CoV-2 infection by sex, age, nationality, and comorbidity, cohorts were exact-matched by these factors to control for their potential impact on our estimates. Given that only 9% of the population of Qatar are $\geq 50$ years of age, our estimates may not be generalizable to other countries where elderly citizens constitute a larger proportion of the total population. |

COVID-19 coronavirus disease 2019, and SARS-CoV-2 severe acute respiratory syndrome coronavirus 2.

**Figure S1.**
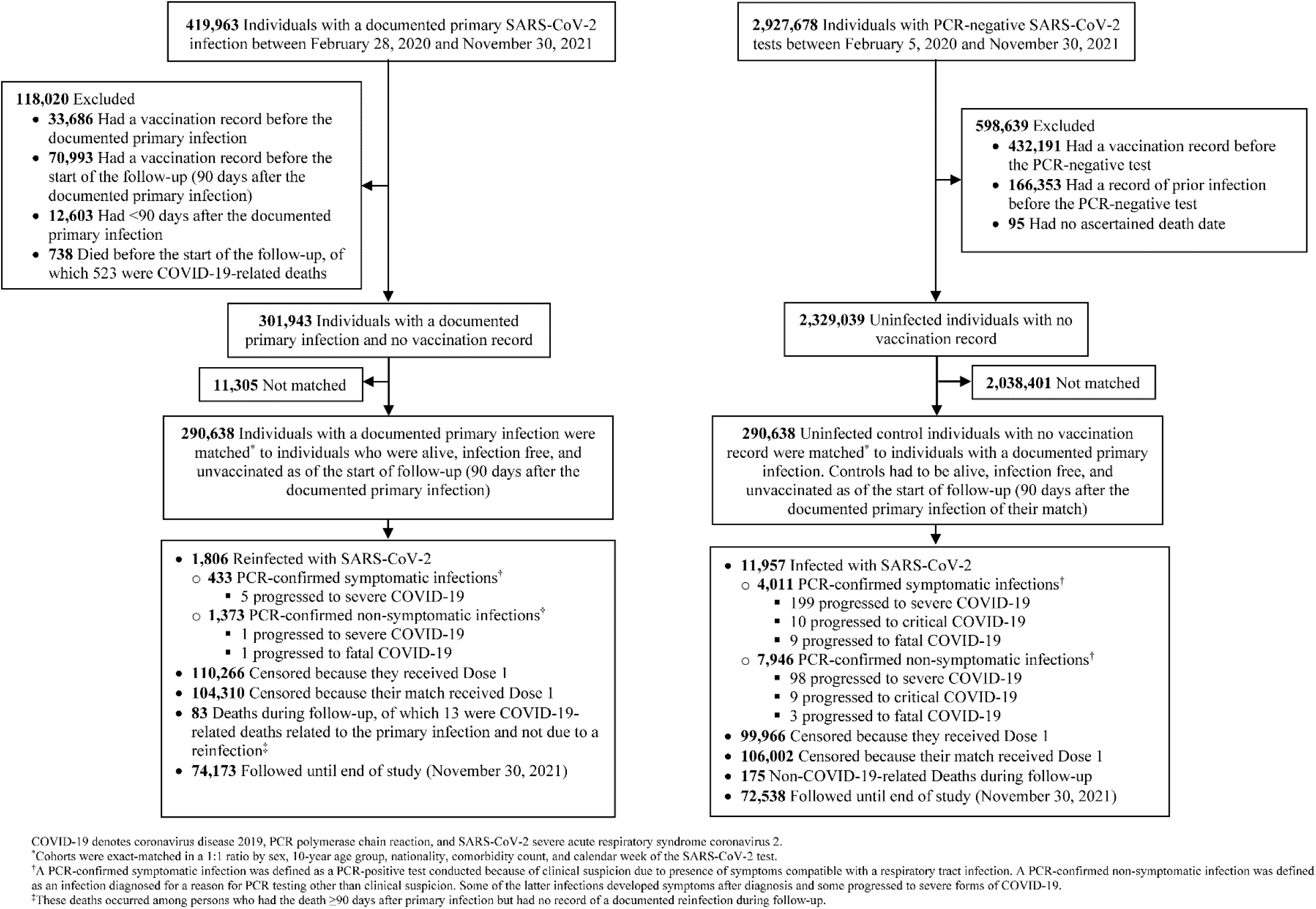
Cohort selection for investigating immune protection of primary infection with a pre-Omicron variant against re- infection with a pre-Omicron variant (Pre-Omicron Reinfection Study).

**Figure S2.**
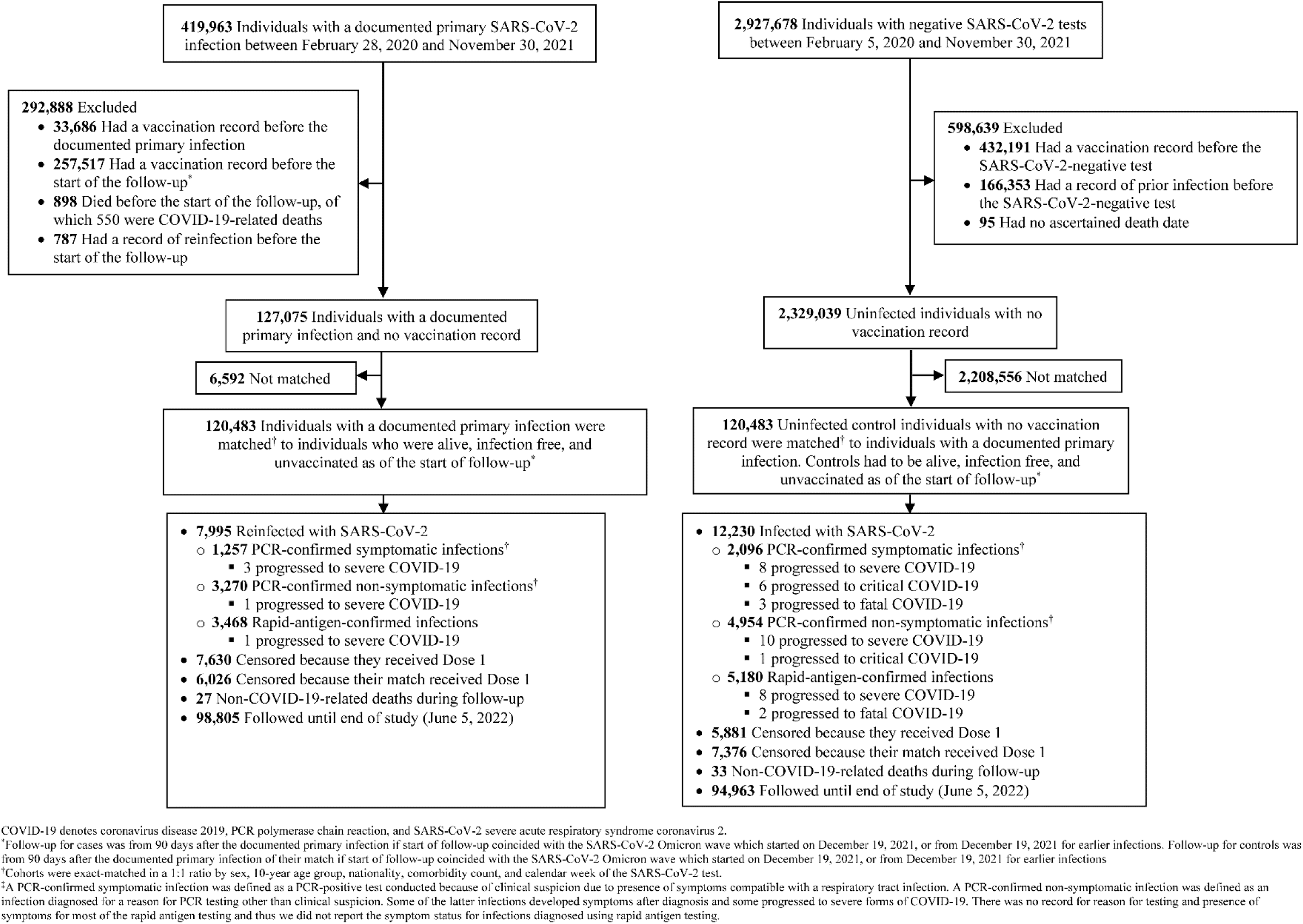
Cohort selection for investigating immune protection of primary infection with a pre-Omicron variant against re- infection with Omicron subvariants (Omicron Reinfection Study).

**Figure S3.**
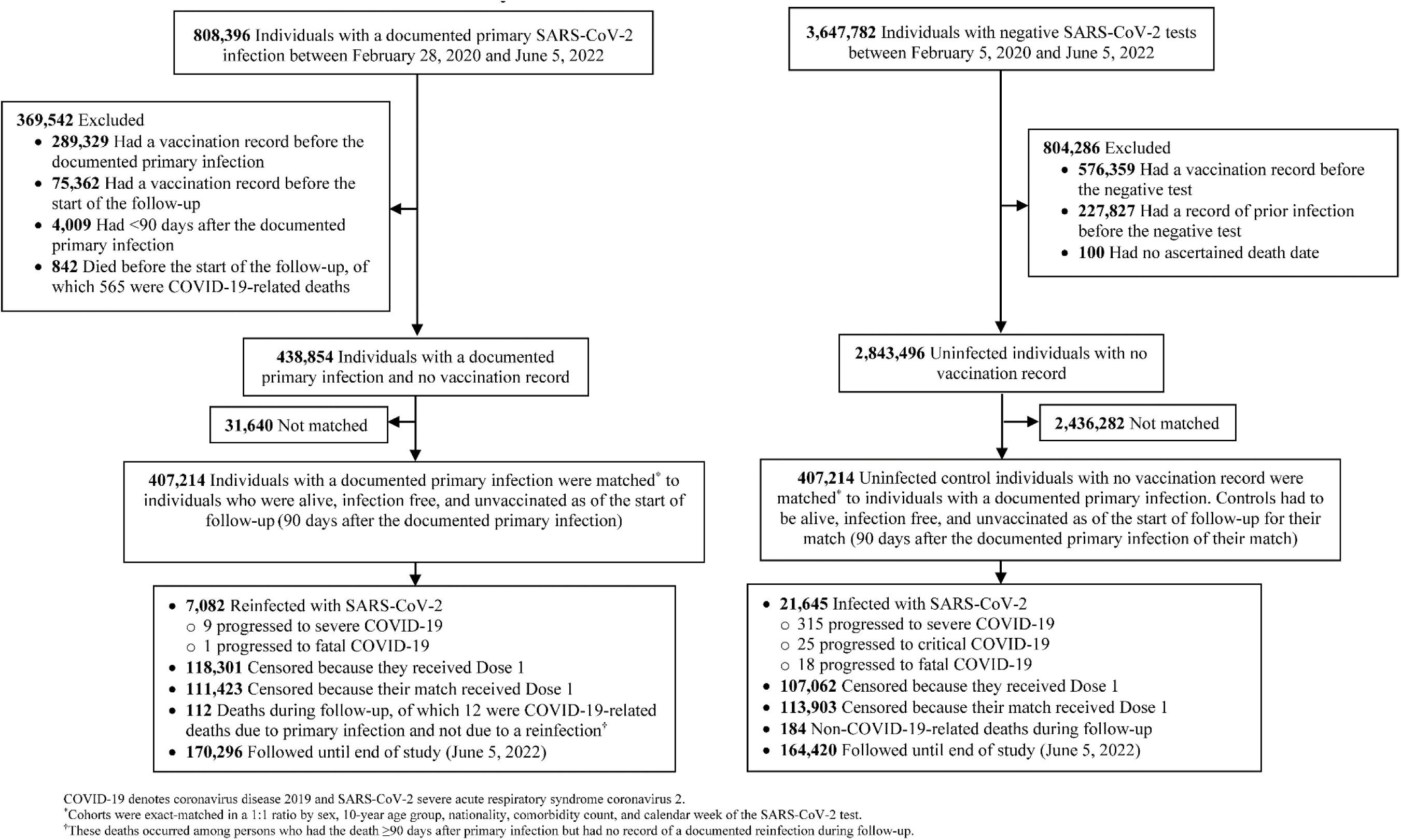
Cohort selection for investigating immune protection of primary infection with any variant against severe, critical, or fatal COVID-19 due to reinfection with any variant.

